# Assessing an ICD-10 Code Approach for Tracking Xylazine-Involved Overdose Deaths in the United States

**DOI:** 10.1101/2024.11.27.24318111

**Authors:** Joseph R. Friedman

**Author notes:** Correspondence to Joseph R. Friedman, MD, PhD, MPH, Department of Psychiatry, University of California, San Diego, 200 W Arbor Dr., San Diego, CA, 92103.

## Abstract

**Introduction:** The national prevalence of the veterinary sedative xylazine in US overdose deaths rose between 2018 and 2021. More updated estimates are limited, partially due to the lack of a dedicated ICD-10 code—a primary mechanism used to specify drugs implicated in overdose deaths in the US, including in the CDC WONDER system, which provides public data requests with a 6-month lag. For other emerging substances lacking dedicated codes, over time umbrella codes have come to *de facto* represent them, yet this has not yet been demonstrated for xylazine.

**Methods:** Overdose deaths in CDC WONDER involving T42.7 (“Antiepileptic and sedative-hypnotic drugs, unspecified”) or T46.5 (“Other antihypertensive drugs, not elsewhere classified”) were compared to two more specific, albeit delayed, sources: NVSS describing national trends in 2018-2021 and SUDORS describing state-level trends in 2020-2022. The CDC WONDER approach was also used to visualize trends in xylazine-involved deaths through Q1 2024 by geography, race/ethnicity, substance co-involvement, and demographic categories.

**Results:** At the national level, concordance between CDC WONDER records and previous NVSS estimates improved after 2019 and became highly similar in 2021 (3,480 vs 3,468 deaths). Concordance was also high for estimates stratified by race, age, and region. At the state-level, across 49 state-year pairs, correlation between CDC WONDER and SUDORS was 0.97. Xylazine-involved deaths doubled between 2021 and 2024 Q1, and racial inequalities widened.

**Discussion:** T42.7 or T46.5, together, may have become the *de-facto* coding scheme representing xylazine-involved deaths. This approach provides more up-to-date results, showing increasing prevalence and worsening racial inequalities in xylazine-involved deaths into 2024.

## Introduction

The widespread proliferation of the veterinary α2-agonist xylazine in the illicit fentanyl supply of the United States (US) has been associated with numerous health risks^1–5^ and labeled an emerging threat by government officials^6^. Previous nationally-representative reports have documented rising xylazine prevalence among overdose deaths between 2018 and 2021^6,7^ and studies drawing on a subset of US states have shown further increases into 2022^8,9^. However, more updated trends are not currently publicly available, partially due to the lack of a dedicated or agreed upon code for xylazine in the tenth revision of the International Classification of Diseases (ICD-10).

ICD-10 codes are a primary mechanism used to track the specific drugs implicated in overdose deaths in the US. In particular they are the main way that substances are specified in the Center for Disease Control and Prevention (CDC) Wide-ranging ONline Data for Epidemiologic Research (WONDER) database^10^, which is used by researchers, policymakers, and the public to access the most up-to-date federal death statistics. CDC WONDER generally makes overdose death statistics available on an approximately 6-month lag, due to inherent delays involved in overdose toxicology testing^11^.

While older drugs such as heroin and cocaine have bespoke ICD-10 codes (T40.1 “Heroin and T40.5 “Cocaine,” respectively), drugs of more recent importance often lack specific codes. However, for some of these substances, such as fentanyl and methamphetamine, over time they have become *de facto* represented by umbrella codes (T40.4 “Other synthetic narcotics” and T43.6 “Psychostimulants with abuse potential”, respectively, in the case of these two substances). These broader codes have become widely understood to mainly represent these drugs of particular importance, although in a small number of cases they may also represent other similar substances^12,13^.

Given the lack of a dedicated ICD-10 code, deaths involving xylazine have largely been tracked using other data sources, such as free text searches in the NVSS system^7^, or the CDC’s State Unintentional Drug Overdose Reporting System (SUDORS)^9^. Both systems leverage more detailed underlying records; however, neither allows the public to use data queries to access continuously updated records, which is a primary advantage of CDC WONDER.

Although there is currently no agreed upon standard set of ICD-10 codes for xylazine, in a recent viewpoint, Gupta et al. used two codes—T42.7 (“Antiepileptic and sedative-hypnotic drugs, unspecified”) or T46.5 (“Other antihypertensive drugs, not elsewhere classified”)—to represent national xylazine deaths^6^. If these codes prove to reliably represent xylazine-involved deaths, there would be numerous potential advantages for producing timely statistics. However, in addition to xylazine, these codes may also represent deaths from other medications, such as levetiracetam, which may bias results^7,8^. Therefore, further assessment is warranted to see how death rates from these ICD-10 codes compare against other more specific, albeit delayed, sources of data.

Additionally, in order to illustrate the possible utility of this method, detailed trends in xylazine-involved deaths through March 2024 are provided by geography, race/ethnicity, substance co-involvement, and demographic categories.

## Methods

The CDC WONDER platform was used to describe overdose deaths involving the T42.7 or T46.5 multiple cause of death ICD-10 codes, from January 2018-March 2024^10^. Overdose deaths were defined as those corresponding to underlying cause of death codes X40–X44 (unintentional), X60–X64 (suicide), X85 (homicide), and Y10–Y14 (undetermined intent). 95% uncertainty intervals were estimated assuming a Poisson distribution. Data from 2023 and 2024 are provisional and may be updated in final numbers. Estimates from January-March (Q1) 2024 were annualized to reflect full year totals should Q1 death rates persist. Race/ethnicity categories were used as defined in the underlying data. See supplemental methods for more details on data preparation.

Xylazine-involved deaths using the ICD-10 code based approach described above were compared to two more specific, albeit delayed, sources describing national and state-level trends in xylazine deaths: NVSS and SUDORS. NVSS data were available from a report published by Spencer et al., which examined free text fields on all death certificates in the system to describe xylazine deaths between 2018 and 2021 by year, race, age, and region^7^. A publicly available SUDORS database was used to assess annual counts of xylazine-involved deaths between 2020 and 2022 for 29 participating states^9^.

This study was deemed exempt from review and informed consent by the UCSD Institutional Review Board as it uses only publicly available, de-identified records.

## Results

At the national level, overdose deaths coded with either T42.7 or T46.5 initially overestimated xylazine deaths according to NVSS in 2018 (with 260 vs 110 deaths, respectively) and 2019 (728 vs 627 deaths) [Figure 1A]. However, the two measures converged considerably in 2020 (1,611 vs 1,499 deaths) and became highly similar in 2021 (3,480 vs 3,468 deaths). Concordance by race, age, and region in 2021 was also high (Figure 1A).

**Figure 1.**
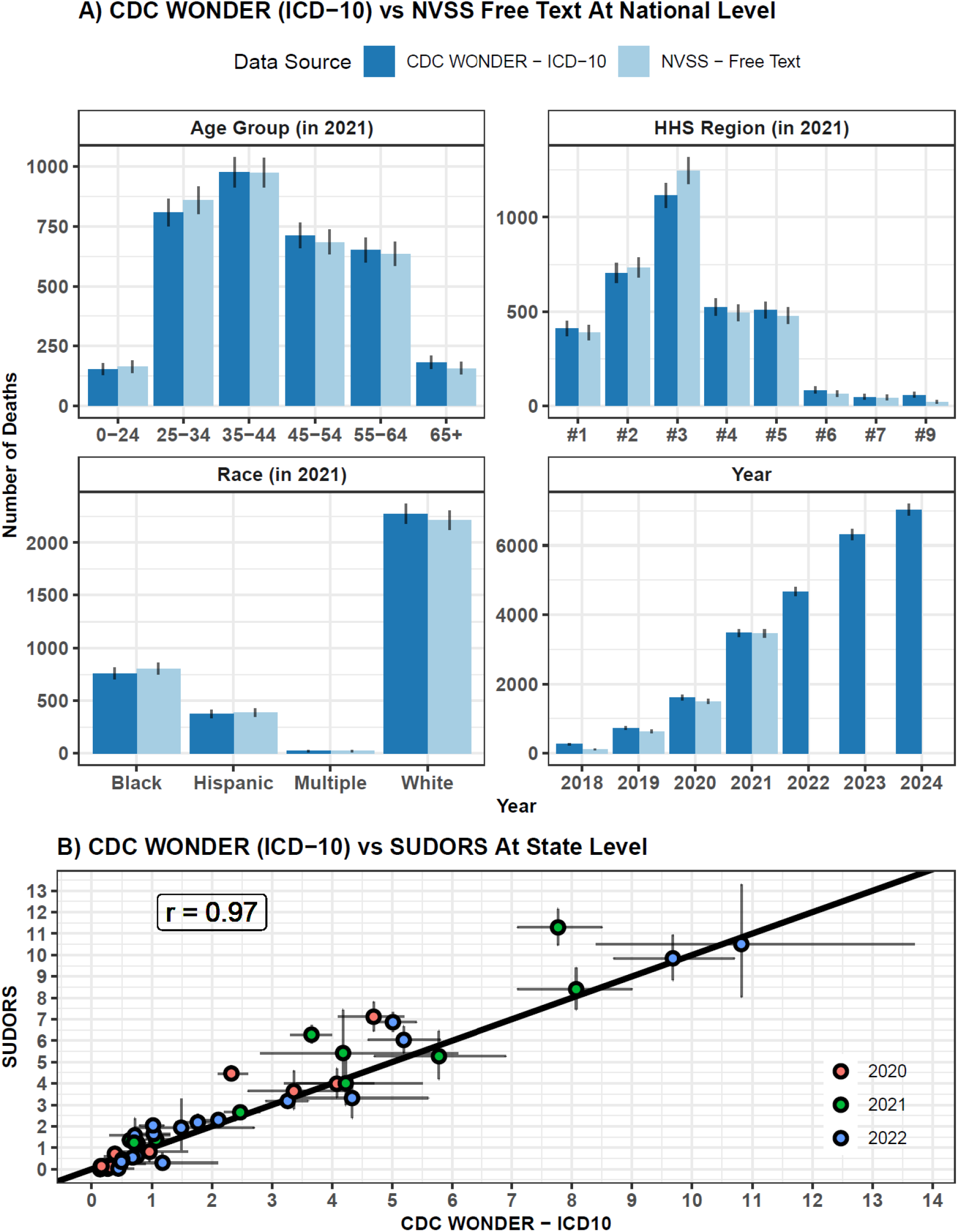
Comparing Xylazine Overdose Deaths Using ICD-10 Codes, SUDORS, and NVSS Free Text. A) Results from the CDC-WONDER ICD-10 code based approach are compared to published results from NVSS by year, race, age group, and HHS region. 95% uncertainty intervals were estimated using a Poisson distribution and are shown with vertical bars. Data from 2023 and 2024 are provisional and may change in final numbers. Numbers from 2024 represent annual equivalents based on data from January-March only. B) State-level estimates are compared between CDC WONDER and SUDORS. Each point represents data from a particular state and year for which data from both sources was available (n=49). SUDORS data are limited to 29 participating jurisdictions. CDC WONDER data are limited to jurisdictions with at least 10 deaths occurring per year. Point color indicates year. 95% uncertainty intervals for both measures are shown with vertical and horizontal bars. Uncertainty intervals for CDC WONDER data were available directl from the platform. Uncertainty intervals for SUDORS data were estimated using a Poisson distribution. A line of equality is shown in black. The overall correlation coefficient across the n=49 observed pairs of datapoints was r=0.97. All CDC WONDER and SUDORS data shown here are finalized (not provisional).

State-level concordance between deaths in CDC WONDER coded with T42.7 and T46.5, and xylazine deaths in the SUDORS system, is shown in Figure 1B. The overall correlation across the n=49 observed state-year pairs was 0.970. Across these 49 paired data points, a regression model predicting CDC WONDER-derived xylazine death rates using SUDORS xylazine death rates as an explanatory variable, with an intercept fixed at 0, showed a slope of 0.854 (95% CI: 0.802 – 0.906), and an adjusted R squared of 0.955. This means that CDC WONDER tended to underestimate trends from SUDORS by about 15%, but the overall predictive power was quite high, with about 95% of variation explained.

Examining the breakdown of deaths from T42.7 and T46.5 shows a shifting pattern over time (Supplemental Figure 1). In 2019 and 2020, the majority of all deaths from either cause were coded as T46.5 “Other antihypertensive drugs, not elsewhere classified”. Deaths identified with this ICD-10 code increased from 112 in 2018, to 460 in 2019, and to 1,208 in 2020, showing a rapid rise, unlikely to be due to increasing deaths from another substance besides xylazine. After 2020, T42.7 “Antiepileptic and sedative-hypnotic drugs, unspecified” became the most common code, and between 2022-2024 it represented over 96% of the total deaths from either code (Supplemental Figure 1).

Results leveraging this ICD-10 code based method are shown in Figure 2 and Supplemental Table 1. The national xylazine-involved overdose death rate increased steadily during the study period and doubled between 2021 (1.05 per 100,000; 95% CI: 1.01-1.08) and Q1 2024 (2.11; 2.06-2.16). Rates in 2023 were higher among men (2.73 per 100,000; 95% CI: 2.65-2.81) than women (1.07; 1.02-1.12), and the highest among the 35-44 (4.06; 3.88-4.26) and 45-54 (3.35; 3.18-3.53) age groups. During the study period the rate among Black and African Americans overtook that of other groups, reaching 3.31 per 100,000 (95%CI: 3.13-3.48) in 2023, representing 1.75 (95%CI: 1.66-1.84) times the national average. In 2023, fentanyl, cocaine, and methamphetamine were co-involved in 96% (95%CI: 93%-98%), 38% (36%-39%) and 23% (22%-24%) of xylazine-involved overdose deaths, respectively. By 2023, all census divisions were affected by xylazine-involved overdose mortality; the Middle Atlantic (5.83 per 100,000; 95% CI: 5.6 - 6.07) and New England (4.48, 4.15 - 4.83), had the highest rates, while the Pacific had the lowest (0.31, 0.26 - 0.36). The states with the highest death rates were Vermont, Pennsylvania, Connecticut, West Virginia, and New York. In 2023, 38 states had at least 10 xylazine-involved overdose death rates, the minimum required for reporting a rate in the CDC WONDER system.

**Figure 2.**
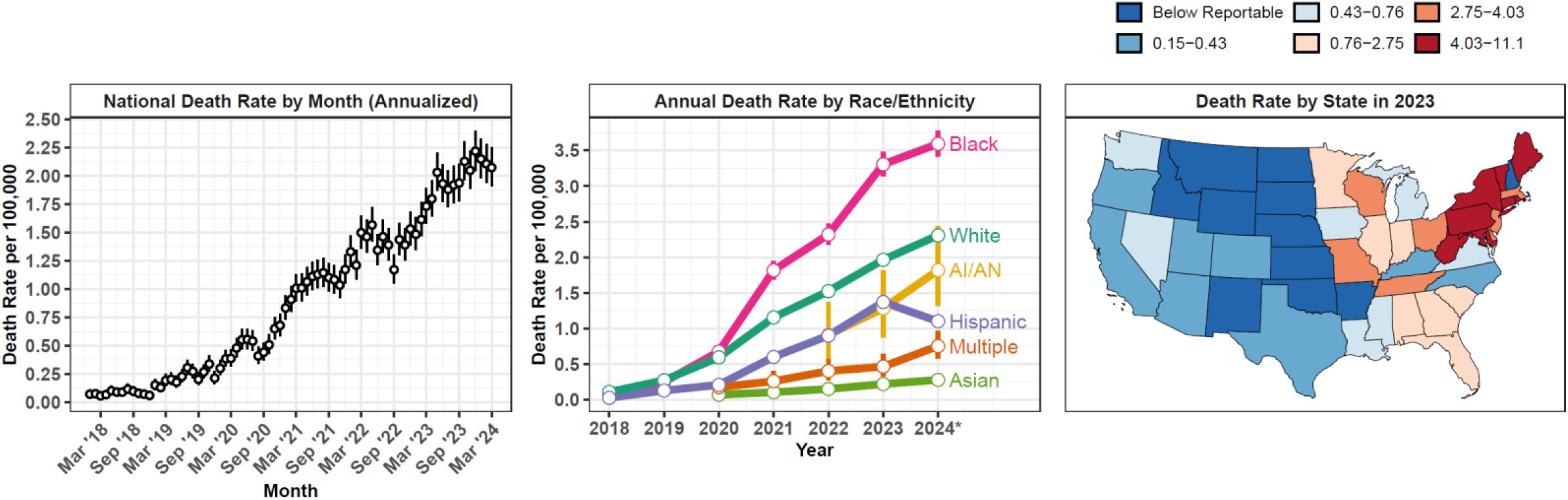
National Estimated Xylazine-lnvolved Overdose Deaths Using CDC Wonder ICD-10 Code Approach. Left. National-level estimates of xylazine-involved overdose deaths per 100,000 are shown by month with 95% uncertainty intervals. Middle. National-level estimates of xylazine-involved overdose deaths per 100,000 are shown by race/ethnicity and year, with 95% uncertainty intervals. Right. Estimates of xylazine-involved overdose deaths per 100,000 in 2023 are shown by state. 95% uncertainty intervals were estimated using a Poisson distribution, as they are not directly available for all estimated quantities in CDC WONDER (e.g. when stratifying data by month). Data from 2023 and 2024 are provisional and may change in final numbers. *Numbers from 2024 shown in the middle panel represent annual equivalents based on data from January-March only.

## Discussion

By 2021, a high concordance was observed at the national level between estimates of xylazine-involved overdose deaths in the CDC WONDER system (using the T42.7 or T46.5 ICD-10 codes) and those of the NVSS system. At the state-level, compared to the SUDORS system, the ICD-10 code based approach somewhat underestimated xylazine-involved death rates, however still showed a high predictive power and correlation. It is important to note that there are underlying differences between CDC WONDER, the NVSS system, and SUDORS, and identical results would not be expected (see supplement). Instead, each of these methods represents a signal from a slightly different set of populations, measured in slightly different ways, yet which should all be roughly comparable in terms of big picture results. Therefore, rather than a formal validation study, this analysis seeks to establish if these various methods of representing xylazine-overdose deaths are roughly compatible—and it concludes that they likely are, at least after 2019.

The breakdown of deaths between these two codes (T42.7 or T46.5) suggests that both were likely initially used to code for xylazine-involved deaths between 2018 and 2021, with T46.7 predominating in 2019 and 2020. After 2021, T42.7 appears to have become the default code for this purpose, with deaths corresponding to T46.5 dropping to very low levels. This suggests that moving forward, using T42.7 as the primary code for xylazine may be sufficient. Nevertheless, historical analyses including data from before 2022 should likely also use T46.5. These results suggest that these two ICD-10 codes, together, may have become the *de-facto* coding scheme used to represent xylazine-involved deaths, similar to other umbrella codes that have taken on a similar role for fentanyl and methamphetamine. This could allow researchers, policymakers, and the public to assess xylazine-involved death trends in CDC WONDER by race, geography, age, sex, and other dimensions, with an approximately 6-month lag. Rather than replacing analyses relying on more detailed systems—such as SUDORS or free text in the NVSS system—the ICD-10 code based approach may simply provide a method of measuring xylazine-involved deaths in a more rapid and publicly transparent fashion.

The small—and likely stable—number of annual deaths represented by these codes involving levetiracetam and other non-xylazine agents appear to have a minimal effect on the total number of deaths after 2019. However, ongoing validation against subsequently released NVSS and SUDORS records would be valuable, to confirm that these data sources continue to be roughly comparable.

Even if a *de facto* set of ICD-10 codes for xylazine is established, numerous challenges remain for tracking xylazine-involved deaths. Given limited testing, the death tolls shown here—using any of the data sources available—almost certainly represent underestimates of the true spread of xylazine^6,7^. Xylazine is also only one of numerous novel substances of concern emerging in the US illicit drug supply, which are spreading rapidly, and lack dedicated ICD-10 codes. Across all of these substances of novel importance, more flexible and transparent systems are needed to track the diverse substances increasingly involved in overdose deaths and provide results to the public in a rapid and transparent fashion.

Leveraging this method also provides insights into recent trends in the spread of xylazine. The prevalence of xylazine-involved overdose deaths has continued to rise sharply in recent years. Although the Northeast remains the most affected region, as xylazine continues to advance from East to West across the nation^14^, it is now present in all major US regions, and the vast majority of states. Racial inequalities in xylazine-involved overdose mortality have also risen sharply—with Black and African Americans almost twice as affected as the national average by 2023—which warrants tailored interventions to reduce inequalities directly^15^.

## Data Availability

All data used in this article are publicly available at wonder.cdc.gov

https://wonder.cdc.gov/

## Supplement

**Supplemental Figure 1.**
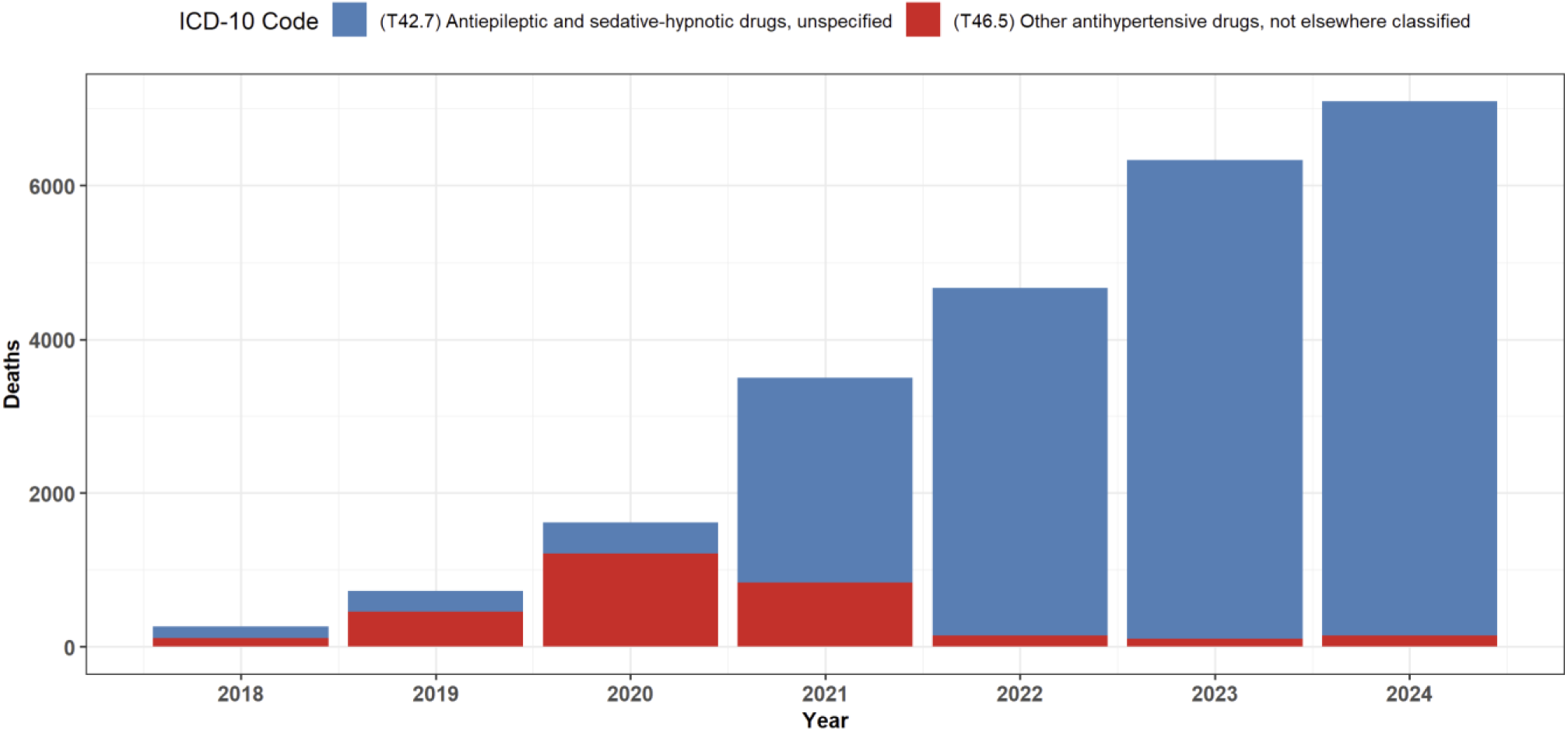
Estimated Deaths from T42.7 vs T46.5. Overdose deaths involving these two ICD-10 codes are shown for 2018-2024. Data from 2023 and 2024 are provisional and may change in final numbers. Numbers from 2024 represent annual equivalents based on data from January-March only.

**Supplemental Table 1.** Key Trends in Estimated Xylazine-involved Overdose Deaths. Estimates of xylazine-involved overdose deaths per 100,000 are shown stratified by several dimensions of interest with 95% uncertainty intervals. Uncertainty was estimated using a Poisson distribution, as it was not directly available for all estimated quantities in CDC WONDER (e.g. when stratifying data by month). Rates were compared to the national average in 2023 to provide a rate ratio for all quantities shown. Data from 2023 and 2024 are provisional and may change in final numbers. *Numbers from 2024 represent annual equivalents based on data from January-March only.

|  | Deaths | Death Rate | Rate Ratio vs. 2023<br>National Average |
| --- | --- | --- | --- |
| **Year** |  |  |  |
| 2018 | 260 | 0.08 (0.07 - 0.09) | 0.04 (0.04 - 0.05) |
| 2019 | 728 | 0.22 (0.21 - 0.24) | 0.12 (0.11 - 0.13) |
| 2020 | 1611 | 0.49 (0.47 - 0.51) | 0.26 (0.25 - 0.27) |
| 2021 | 3480 | 1.05 (1.01 - 1.08) | 0.55 (0.54 - 0.57) |
| 2022 | 4662 | 1.4 (1.36 - 1.44) | 0.74 (0.72 - 0.76) |
| 2023 | 6309 | 1.89 (1.85 - 1.94) | 1.0 |
| 2024* | 7032* | 2.11 (2.06 - 2.16) | 1.11 (1.09 - 1.14) |
| **Gender in 2023** |  |  |  |
| Female | 1803 | 1.07 (1.02 - 1.12) | 0.57 (0.54 - 0.59) |
| Male | 4506 | 2.73 (2.65 - 2.81) | 1.44 (1.4 - 1.48) |
| **Age Group in 2023** |  |  |  |
| 15-24 years | 253 | 0.57 (0.5 - 0.65) | 0.3 (0.27 - 0.34) |
| 25-34 years | 1290 | 2.84 (2.68 - 2.99) | 1.5 (1.42 - 1.58) |
| 35-44 years | 1776 | 4.06 (3.88 - 4.26) | 2.15 (2.05 - 2.25) |
| 45-54 years | 1355 | 3.35 (3.18 - 3.53) | 1.77 (1.68 - 1.87) |
| 55-64 years | 1169 | 2.78 (2.62 - 2.94) | 1.47 (1.38 - 1.55) |
| 65-74 years | 398 | 1.18 (1.07 - 1.3) | 0.62 (0.56 - 0.69) |
| 75-84 years | 41 | 0.23 (0.17 - 0.32) | 0.12 (0.09 - 0.17) |
| **Race/Ethnicity in 2023** |  |  |  |
| AI/AN | 31 | 1.28 (0.87 - 1.82) | 0.68 (0.46 - 0.96) |
| Asian, NH | 44 | 0.22 (0.16 - 0.29) | 0.11 (0.08 - 0.15) |
| Black, NH | 1391 | 3.31 (3.13 - 3.48) | 1.75 (1.66 - 1.84) |
| White, NH | 3854 | 1.96 (1.9 - 2.03) | 1.04 (1.01 - 1.07) |
| Multiple, NH | 37 | 0.46 (0.33 - 0.64) | 0.24 (0.17 - 0.34) |
| Hispanic | 872 | 1.37 (1.28 - 1.46) | 0.72 (0.68 - 0.77) |
| **Census Division in 2023** |  |  |  |
| New England | 678 | 4.48 (4.15 - 4.83) | 2.37 (2.19 - 2.55) |
| Middle Atlantic | 2444 | 5.83 (5.6 - 6.07) | 3.08 (2.96 - 3.21) |
| East North Central | 1060 | 2.25 (2.12 - 2.39) | 1.19 (1.12 - 1.26) |
| West North Central | 254 | 1.17 (1.03 - 1.32) | 0.62 (0.54 - 0.7) |
| South Atlantic | 1226 | 1.82 (1.72 - 1.92) | 0.96 (0.91 - 1.02) |
| East South Central | 323 | 1.65 (1.47 - 1.84) | 0.87 (0.78 - 0.97) |
| West South Central | 86 | 0.21 (0.17 - 0.25) | 0.11 (0.09 - 0.13) |
| Mountain | 73 | 0.29 (0.22 - 0.36) | 0.15 (0.12 - 0.19) |
| Pacific | 165 | 0.31 (0.26 - 0.36) | 0.16 (0.14 - 0.19) |
| **Top Ten Most Affected States in 2023** |  |  |  |
| Vermont | 72 | 11.13 (8.71 - ) | 5.88 (4.6 - 7.4) |
| Pennsylvania | 1037 | 7.99 (7.51 - 8.5) | 4.22 (3.97 - 4.49) |
| Connecticut | 278 | 7.67 (6.79 - 8.62) | 4.05 (3.59 - 4.56) |
| West Virginia | 123 | 6.93 (5.76 - 8.27) | 3.66 (3.04 - 4.37) |
| New York | 1045 | 5.31 (4.99 - 5.64) | 2.81 (2.64 - 2.98) |
| Maine | 67 | 4.84 (3.75 - 6.14) | 2.55 (1.98 - 3.24) |
| Maryland | 259 | 4.2 (3.71 - 4.75) | 2.22 (1.96 - 2.51) |
| Rhode Island | 45 | 4.11 (3 - 5.51) | 2.17 (1.59 - 2.91) |
| New Jersey | 362 | 3.91 (3.52 - 4.33) | 2.06 (1.86 - 2.29) |
| Wisconsin | 225 | 3.82 (3.34 - 4.35) | 2.02 (1.76 - 2.3) |
| **Substances Co-Involved in 2023** |  |  |  |
| Fentanyl | 6028 | 1.81 (1.76 - 1.85) | 0.96 (0.93 - 0.98) |
| Cocaine | 2375 | 0.71 (0.68 - 0.74) | 0.38 (0.36 - 0.39) |
| Methamphetamine | 1431 | 0.43 (0.41 - 0.45) | 0.23 (0.22 - 0.24) |
| Benzodiazepine | 982 | 0.29 (0.28 - 0.31) | 0.16 (0.15 - 0.17) |
| Heroin | 644 | 0.19 (0.18 - 0.21) | 0.1 (0.09 - 0.11) |

### Steps Taken to Conduct the Analysis

1. **NVSS Data:** Data describing national counts of xylazine-involved deaths from 2018 to 2021 from the National Vital Statistics System were accessed from the published tables of the following report: “Spencer M, Cisewski J, Warner M, Garnett M. Drug Overdose Deaths Involving Xylazine, United States, 2018–2021. National Center for Health Statistics (U.S.); 2023. doi:10.15620/cdc:129519”. This analysis looked at overdose deaths, as indicated by the standard set of underlying cause of death codes for this purpose, including X40–X44 (unintentional), X60– X64 (suicide), X85 (homicide), and Y10–Y14 (undetermined intent). This report leveraged a free text search approach drawing on the live, dynamically updated NVSS database. This database is also abstracted to create the death counts in the CDC WONDER database, however with fewer details. The underlying NVSS database includes free text from original death certificates, which were systematically searched in this analysis to yield death counts by year from 2018 to 2021, as well as race, age group, and HHS region. Given that the concordance was noted to improve over time in year-to-year results, for the other examined dimensions, the latest year of available data (2021) was used for comparative purposes.
2. **SUDORS Data:** Data describing state-year specific counts of xylazine-involved deaths were downloaded from the public data download provided on the SUDORS data dashboard: “Centers for Disease Control and Prevention. State Unintentional Drug Overdose Reporting System (SUDORS). Final Data. Atlanta, GA: US Department of Health and Human Services, CDC; [October 18^th^, 2024]. Access at: https://www.cdc.gov/overdose-prevention/data-research/facts-stats/sudors-dashboard-fatal-overdose-data.html“. SUDORS is a separate data system for participating states to upload overdose data, with a greater level of detail compared to the CDC WONDER system. Data describing xylazine-involved overdoses were available in the ‘final’ records, for 2020 through 2022, by state.
3. **CDC WONDER Data:** Data matching the format of the aforementioned two sources were obtained from the CDC WONDER platform (https://wonder.cdc.gov/) for finalized and provisional data from 2018 through the latest available records. Overdose deaths were defined as those corresponding to underlying cause of death codes X40–X44 (unintentional), X60–X64 (suicide), X85 (homicide), and Y10–Y14 (undetermined intent). Deaths were selected when they also contained either multiple cause of death code T42.7 (“Antiepileptic and sedative-hypnotic drugs, unspecified”) or T46.5 (“Other antihypertensive drugs, not elsewhere classified”). Data from 2018-2022 are finalized and will not change. Data from 2023 and 2024 are provisional and may change in final numbers. Numbers from 2024 represent annual equivalents based on data from January-March only. To calculate this, numbers from Q1 of 2024, January – March 2024 were accessed, and multiplied by 4 to estimate annual totals. Population numbers were obtained from CDC WONDER for calculating per-capita rates.
4. **Uncertainty Intervals:** 95% uncertainty intervals were estimated for all numbers shown in the main text figures. For CDC WONDER based death rates shown in figure 2, uncertainty intervals were available directly from the CDC WONDER platform. For all other count data, 95% uncertainty intervals were estimated using a Poisson distribution, using the exact method. This was accomplished in R using the qchisq() function.
5. **Correlation Coefficients and linear models:** correlation coefficients and linear models were assessed in R using the cor() function and lm() function.
6. **Data visualization:** data were visualized using the ggplot package in R.

### Methodological Considerations

Arguably, the 3 data sources compared in this analysis all represent “signals” of the same underlying process (the national spread of xylazine), however important differences do exist between them. This analysis relies on the premise that these differences do not undermine the approximate comparability between each data source, however they do inform why numbers vary to a small degree from one source to another. Below is a non-exhaustive list of some of the differences between the data sources.

1. The NVSS database used in the Spencer et al. report is continuously updated, even after “finalized” records are abstracted from it to populate CDC WONDER. They may differ from other published provisional or final overdose statistics.
2. SUDORS is a separate underlying data system, and is limited to a select set of participating states. Therefore, it cannot be used for head-to-head national-level comparisons. Additionally, counts from the SUDORS system reflect a more limited set of underlying cause of death codes, only corresponding to unintentional or undetermined intent overdoses. The small number of overdose deaths coded as corresponding to suicides or homicides are not included here but are included for the CDC WONDER and NVSS data sources.
3. Data in NVSS and SUDORS are specifically coded for xylazine by name. Data in CDC WONDER coded with T42.7 or T46.5reflect xylazine deaths as well as a small number of deaths from other substances, including the anti-convulsant levetiracetam. Additionally, other emerging α2-agonists, such as medetomidine, are likely to be captured using these same ICD-10 codes. Although currently xylazine represents the vast majority of α2-agonist-involved deaths, this may change in the future, requiring additional code specifications. See: “Palamar JJ, Krotulski AJ. Medetomidine Infiltrates the US Illicit Opioid Market. JAMA. Published online September 4, 2024. doi:10.1001/jama.2024.15992”

